# Advancing Post-Traumatic Seizure Classification and Biomarker Identification: Information Decomposition Based Multimodal Fusion and Explainable Machine Learning with Missing Neuroimaging Data

**DOI:** 10.1101/2022.10.22.22281402

**Authors:** Md Navid Akbar, Sebastian F. Ruf, Ashutosh Singh, Razieh Faghihpirayesh, Rachael Garner, Alexis Bennett, Celina Alba, Marianna La Rocca, Tales Imbiriba, Deniz Erdoğmuş, Dominique Duncan

## Abstract

A late post-traumatic seizure (LPTS), a consequence of traumatic brain injury (TBI), can potentially evolve into a lifelong condition known as post-traumatic epilepsy (PTE). Presently, the mechanism that triggers epileptogenesis in TBI patients remains elusive, inspiring the epilepsy community to devise ways to predict which TBI patients will develop PTE and to identify potential biomarkers. In response to this need, our study collected comprehensive, longitudinal multimodal data from 48 TBI patients across multiple participating institutions. A supervised binary classification task was created, contrasting data from LPTS patients with those without LPTS. To accommodate missing modalities in some subjects, we took a two-pronged approach. Firstly, we extended a graphical model-based Bayesian estimator to directly classify subjects with incomplete modality. Secondly, we explored conventional imputation techniques. The imputed multimodal information was then combined, following several fusion and dimensionality reduction techniques found in the literature, and subsequently fitted to a kernel- or a tree-based classifier. For this fusion, we proposed two new algorithms: recursive elimination of correlated components (RECC) that filters information based on the correlation between the already selected features, and information decomposition and selective fusion (IDSF), which effectively recombines information from decomposed multimodal features. Our cross-validation findings showed that the proposed IDSF algorithm delivers superior performance based on the area under the curve (AUC) score. Ultimately, after rigorous statistical comparisons and interpretable machine learning examination using Shapley values of the most frequently selected features, we recommend the two following magnetic resonance imaging (MRI) abnormalities as potential biomarkers: the left anterior limb of internal capsule in diffusion MRI (dMRI), and the right middle temporal gyrus in functional MRI (fMRI).

## 1. Introduction

### 1.1 Background

Post-traumatic epilepsy (PTE) constitutes a subtype of acquired epilepsy, originating from a traumatic brain injury (TBI) inflicted by an external force, such as an accident or a fall. A notable risk factor for PTE is the incidence of a post-traumatic seizure (PTS), particularly a late PTS (LPTS), which transpires a minimum of one week after the injury. (Gugger and Diaz-Arrastia, 2022). Presently, there is a deficiency of efficient pharmacological interventions capable of preventing PTE seizures, underlining the necessity for researchers in the field of epilepsy to identify potential biomarkers and devise methods to predict the likelihood of TBI patients developing LPTS (Piccenna et al., 2017). TBI causes extensive damage to both functional and structural features. Therefore, to comprehend the effects of TBI comprehensively, a multimodal approach is essential. (Sharp et al., 2011). The Epilepsy Bioinformatics Study for Antiepileptogenic Therapy (EpiBioS4Rx) (Vespa et al., 2019) is at the forefront of this initiative, aiming to conceptualize and conduct preclinical trials of antiepileptogenic therapies, leveraging multimodal information. This lays the groundwork for future planning of clinical trials.

### 1.2 Related Work

#### Multimodal Fusion

For classification tasks utilizing multiple modality information, three major approaches are found in the literature. They are, namely: late, intermediate, and early fusion. Late fusion involves combining predictions from multiple different base learners (Base), or heterogeneous ensemble classification, and it has been shown to be an efficient way of improving predictive accuracy in machine learning (ML) (Wolpert, 1992). As a form of late fusion, stacking involves learning how to best combine the classifiers, in addition to combination using simple statistics, such as voting or averaging (Džeroski and Ženko, 2004). Džeroski et al. claimed that heterogeneous base learners, stacked with a final meta classifier, can perform comparably, if not better, to the best of the individual classifiers (Džeroski and Ženko, 2004). In the second major approach, intermediate fusion, features or information extracted from the individual modalities are aggregated, and subsequently fed to a single classifier. Since fusion in this method occurs in the feature space, it is often more interpretable and also allows the application of multivariate information theoretic fusion (Brown, 2009). The final approach, early fusion, involves representing the raw individual modality information collectively in a common space. From this aggregated information, an ML pipeline can then extract relevant features and perform the desired classification. Early fusion can be challenging to implement, since meaningful aggregation of different types of modalities (e.g. spatial and temporal) is not always straightforward (Zhang et al., 2020). Past studies involving magnetic resonance imaging (MRI) and electroencephalography (EEG) have found intermediate and late fusion to be less affected by noise, misregistration, and to have lower complexity; compared to early fusion (Lewis et al., 2007; Zhang et al., 2020).

#### Missing Data

While acquiring data from all modalities for each subject is ideally desirable, it is often not feasible. This creates a challenge of handling such missing modality information. Missing data has historically been dealt with imputation: using the existing data to make estimates for the missing values. Imputation techniques generally take either the form of a univariate approach that considers each missing feature/ modality in isolation across all subjects (such as mean imputation), or a multivariate approach that considers the other available features/ modalities, which could be within a subject or across all subjects (such as k-nearest neighbor, kNN imputation (Scheffer, 2002)). Following imputation, standard fusion and classification pipelines can be implemented. Aside from conventional imputation, contemporary approaches include jointly performing data imputation and self-representation learning (Liu et al., 2021), or graphical model estimators that marginalize missing attributes (Murphy, 2012).

#### Seizure Classification

TBI causes widespread damage to both functional and structural features. Since each imaging modality tends to pick up only certain items from this entire set of features, it is essential to consider multiple modalities for such classification (Sharp et al., 2011). Whereas many studies have investigated a unimodal strategy for seizure classification, few have involved a multimodal approach. Two such studies explored non-TBI seizure classification from EEG and structural MRI (Memarian et al., 2015), and from EEG and electrocardiography (ECG) (Yang et al., 2022). Akrami et al. examined PTE classification, utilizing functional MRI (fMRI) and T2-weighted fluid-attenuated inversion recovery (T2-FLAIR) for lesion information (Akrami et al., 2021). However, to our knowledge, no earlier work investigated the classification of LPTS from an incomplete multimodal dataset, proposed suitable novel fusion algorithms, and searched for modality-specific potential biomarkers. Aside, the current work builds on our earlier preliminary works in preprocessing and analyzing the data (Akbar et al., 2020, 2021; Faghihpirayesh et al., 2021) from the ongoing multi-center EpiBioS4Rx cohort.

### 1.3 Contributions

In the current study, we initiate a binary classification task to distinguish between subjects experiencing LPTS and those who do not, in selected subjects from the EpiBioS4Rx cohort. We elaborate on our data collection and preparation strategies in Section 2, for diffusion-weighted MRI (dMRI), EEG, and fMRI. While making meaningful inferences involving multimodal fusion is a challenging problem, it is complicated further by the absence of information about certain modalities in certain subjects. This work attempts to address that missing data challenge as well. Furthermore, we strive to identify potential biomarkers within each modality. To encapsulate our unique contributions:

1. We pioneer a binary LPTS classification task leveraging a multimodal dataset of dMRI, EEG, and fMRI. Additionally, we incorporated lesion information from T2-FLAIR during the fMRI preprocessing.
2. In response to missing modality in certain subjects, we extend a graphical model-based Bayesian estimator for direct classification using the incomplete original dataset. Simultaneously, we explore various univariate and multivariate imputation techniques to create imputed datasets.
3. We implement several techniques from intermediate and late fusion for integrating multimodal information from the imputed datasets. Regarding intermediate fusion, we introduce two novel algorithms: one that filters multivariate information based on correlation between the previously selected features/ components, and another that selectively fuses information following the decomposition of the available multivariate information.
4. Lastly, we identify modality-specific potential biomarkers. This is achieved through statistical comparisons and interpretable machine learning analyses (using Shapley values) of the most frequently selected features distinguishing the LPTS groups.

The structure of the remainder of the paper is as follows: Section 3 presents our empirical findings, Section 4 summarizes our discussion, and Section 5 offers conclusions drawn from our work.

## 2. Methods

### 2.1 Data Collection

#### 2.1.1 Acquisition

This work was approved by the UCLA Institutional Review Board (IRB#16 − 001576) and the local review boards at each EpiBioS4Rx Study Group institution. Written informed consent to participate in this study was provided by the participants’ legal guardian/next of kin.

According to the EpiBioS4Rx protocol (Vespa et al., 2019), moderate-to-severe TBI subjects with frontal and/or temporal hemorrhagic contusion and Glasgow Coma Scale (GCS, severity of injury, lower is severer) score between 3 − 13 were eligible for enrollment. A total of 48 subjects (12 female, 36 male; age = 42.1 ± 19.3 years; GCS = 7.7 ± 4.4), enrolled in 8 different sites, were chosen for this work. Of these subjects, 17(35%) experienced at least one LPTS, whereas the remaining 31 did not experience any for the entire two-year follow-up period, which is the minimum duration needed to identify about 80 − 90% of the subjects who will eventually develop PTE (Temkin, 2009). Even though the modalities of interest for this work are dMRI, EEG, and fMRI, the acquired data from the subjects included other MRI sequences such as T1-weighted, T2-weighted, T2-FLAIR, etc. The following three sections will summarize the preprocessing steps involved in preparing our three data modalities of interest for the extraction of relevant features.

#### 2.1.2 dMRI Preprocessing

dMRI scans corresponding to multiple diffusion gradient values and directions were collected. Acquired dMRI data was processed in the Oxford FMRIB Software Library (FSL) (Jenkinson et al., 2012), to estimate diffusion tensor imaging (DTI) parameters, such as fractional anisotropy (FA) maps, in subject-specific spaces. Since FA features have been reported earlier to be promising in characterizing PTS (Gupta et al., 2005), we focused on extracting and utilizing FA features in this work.

The collected individual subject FA images were then transformed to a common Montreal Neurological Institute (MNI) space by registering to a standard HCP 1065 DTI FA template, following Pipeline 1 in (Akbar et al., 2021). Finally, tract-based spatial statistic (Smith et al., 2006) of each registered image was carried out using the mask and distance map obtained from the standard template, to extract mean FA values along 63 white matter (WM) tracts and bundles obtained from the JHU-DTI atlas (Zhang et al., 2010). These mean FA values are recorded as the dMRI features (*x*_*d*_).

##### Exclusion Criteria

Of the total 48 subjects in our study, 45 had valid dMRI scans for analysis. From these 45 subjects, we further excluded 4 as they did not meet the two-year followup requirement stipulated by (Temkin, 2009). This left us with a final sample of 41 subjects. Within this group, 14 subjects (representing 34% of the sample) developed LPTS, while the remaining 27 subjects (representing 66% of the sample) did not.

#### 2.1.3 EEG Preprocessing

Epileptiform abnormalities (EAs) such as seizures, periodic discharges (PDs), and abnormal rhythmic delta activity (ARDA), are potential biomarkers of epileptogenesis (Vespa et al., 2016; Faghihpirayesh et al., 2021). Kim et al. (Kim et al., 2018) showed that the presence of EAs in the EEG signal during the acute period following TBI independently predicted PTE in the first year post-injury. Clinicians annotated the obtained EEG to denote the presence of three such EAs (Seizure, PD, and ARDA), which formed our EEG features (*x*_*e*_).

*Exclusion Criteria:* Of the total 48 subjects in our study, 10 had valid EEG recordings (following review by EEG experts) for analysis. All subjects met the two-year follow-up requirement stipulated by (Temkin, 2009). Within this group, 7 subjects (representing 70% of the sample) developed LPTS, while the remaining 3 subjects (representing 30% of the sample) did not.

#### 2.1.4 fMRI Preprocessing

To incorporate lesion information in the rs-fMRI preprocessing, we manually segmented damaged brain tissue into parenchymal contusions and brain edema with ITK-SNAP (Yushkevich et al., 2006) from the acquired 3D T2-FLAIR scans. Brain contusion was defined as a lesion with abnormal signal intensity and hemorrhagic volume > 1 ml, and brain edema was defined as a region surrounding or in the proximity of the contusion with hyperintense signal compared with the WM signal on T2-FLAIR images (Chang et al., 2006). Each segmentation was carried out by a student research assistant and reviewed for clinical accuracy by one of two medical physicians with research experience in neuroradiology (Bennett et al., 2023). For each TBI subject, we obtained a 3D lesion mask including contusion and edema. For this work, the lesion incorporation served to obtain a more accurate preprocessing pipeline, in which brain alterations related to TBI were not mislabeled as brain tissues such as WM, cerebrospinal fluid (CSF), and grey matter (GM). Therefore, we did not consider the edema and contusion separately within each 3D mask. Then, to use the lesion masks in the rs-fMRI preprocessing pipelines, we performed affine registration on each mask using the MNI 152 template with the Linear Registration Tool (FLIRT) of FSL (Jenkinson et al., 2012). Affine transformations were used because they allowed us to maintain the morphological characteristics of the lesions better while at the same time obtaining robust registrations.

The preprocessing used a modification of the unified segmentation normalization of SPM12 (Ashburner and Friston, 2005) as accessed through the CONN toolbox (Whitfield-Gabrieli and Nieto-Castanon, 2012). The literature on normalization suggests that SPM12’s unified segmentation normalization outperforms other pipelines when applied to simulated lesions (Crinion et al., 2007). However, further work (Andersen et al., 2010) showed that to achieve the best performance, information about the lesion should be included, motivating the inclusion of the lesion mask in preprocessing.

The fMRI preprocessing pipeline incorporates information about the lesion by performing SPM12’s unified segmentation normalization using a lesion-modified tissue probability map (TPM). As discussed in (Andersen et al., 2010), modifying the TPM performs implicit cost function normalization because it ultimately causes SPM to ignore the lesion areas during normalization. The TPM normally provides a prior probability of a given voxel belonging to one of 6 tissue classes; GM, WM, CSF, skull, soft tissue, and other (Ashburner, 2009). SPM12’s default TPM was augmented for each subject with a 7th tissue class corresponding to the MNI lesion mask for that subject, thereby setting the prior probability of GM, WM, or CSF in the lesion areas to 0. The modified TPM was created by a new function in the CONN toolbox, conn createtpm. Finally, using the AAL3 atlas (Rolls et al., 2020), the means of the positive (*x*_*f* −*p*_) and negative (*x*_*f* −*n*_) parcel-to-parcel correlations (Faria et al., 2012), and the mean of the lesion volume overlap with parcellations (*x*_*f* −*o*_) were recorded as the fMRI features.

*Exclusion Criteria:* Of the total 48 subjects in our study, 36 had valid fMRI scans for analysis. From these 36 subjects, we further excluded 4 as they did not meet the two-year followup requirement stipulated by (Temkin, 2009). This left us with a final sample of 32 subjects. Within this group, 12 subjects (representing 38% of the sample) developed LPTS, while the remaining 20 subjects (representing 62% of the sample) did not.

### 2.2 Model Development

#### 2.2.1 Data Partitions

While the goal of the study was to acquire all three modalities of interest (*x*_*d*_, *x*_*e*_, *x* _*f*_) for each subject, all subjects did not have all modalities. Table 1 shows the number of subjects for each combination of modalities, as well as the relative number of subjects within each modality configuration. Out of the included total *N* = 48 subjects, only seven had information available from all three modalities. Of the other 41 subjects, 21 had a single missing modality, while the remaining 20 had two missing modalities.

**Table 1.** Distribution of subjects by demographics, injury severity (lower is severer), available modalities, and LPTS labels.

|  | Subjects | Female | Male | Age | GCS | fMRI | dMRI | EEG | LPTS | No LPTS |
| --- | --- | --- | --- | --- | --- | --- | --- | --- | --- | --- |
|  | 5 | 1 | 4 | 41.0±22.1 | 6.6±5.0 | ✓ |  |  | 1 | 4 |
|  | 13 | 2 | 11 | 41.7±14.8 | 7.4±4.4 |  | ✓ |  | 2 | 11 |
|  | 2 | 0 | 2 | 41.0±19.8 | 3.0±0.0 |  |  | ✓ | 2 | 0 |
|  | 20 | 7 | 13 | 46.1±22.1 | 7.7±4.5 | ✓ | ✓ |  | 7 | 13 |
|  | 1 | 0 | 1 | 67.0±0.0 | 9.0±0.0 |  | ✓ | ✓ | 1 | 0 |
|  | 7 | 2 | 5 | 29.0±13.4 | 10.1±4.5 | ✓ | ✓ | ✓ | 4 | 3 |
| Total: | 48 | 12 | 36 | 42.1±19.3 | 7.7±4.4 | 32 | 41 | 10 | 17 | 31 |

From the total *N* subjects, we then created train and test partitions, following a five-fold cross-validation (CV). Any missing modalities were handled using methods described in Section 2.2.3. We concatenated all *D* features (166 × 3 for the three types of fMRI, 63 for dMRI, 3 for EEG) along the columns of 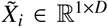, for *I* ∈ *n* samples in each CV train fold. Each 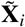, representing an individual subject, is then stacked vertically as rows to give the training partition (**X**_train_) for each CV round

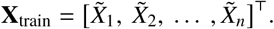

Each test partition (**X**_test_) is constructed in a similar manner with *m* = *N* − *n* samples. The complete feature set, including the train and the test sets, is denoted by [**X** = **X**_train_, **X**_test_]^T^, where **X** ∈ ℝ^*N*×*D*^. Similarly, the train-test partitions for the complete set of LPTS labels **y** ∈ ℝ^*N*×1^, corresponding to any CV round, could be expressed as **y** = [**y**_train_, **y**_test_]^T^.

#### 2.2.2 Classification Framework

The comprehensive modeling framework employed for the LPTS classification task is illustrated in Fig. 1. To begin, highdimensional features drawn from diverse modalities, symbolized as **x**_*d*_, **x**_*e*_, **x** _*f*_, are concatenated. These combined features then pass through an imputation stage, which estimates missing data points using the available data. The data, now filled in, proceed to the dimension reduction stage, which projects the incoming high-dimensional data into a more tractable lower-dimensional space. This reduced dataset is subsequently fit to a classification stage that predicts the binary seizure label, denoted as ŷ. An all-encompassing fusion stage determines the fusion technique to be utilized at any pertinent stage of the pipeline. The *(Yes*/ *No)* choices indicate whether a stage is active or not: if ‘yes’, the stage employs one of several techniques discussed in subsequent sections; if ‘no’, the stage becomes a passive conduit, with its output remaining identical to its input.

**Fig. 1.**
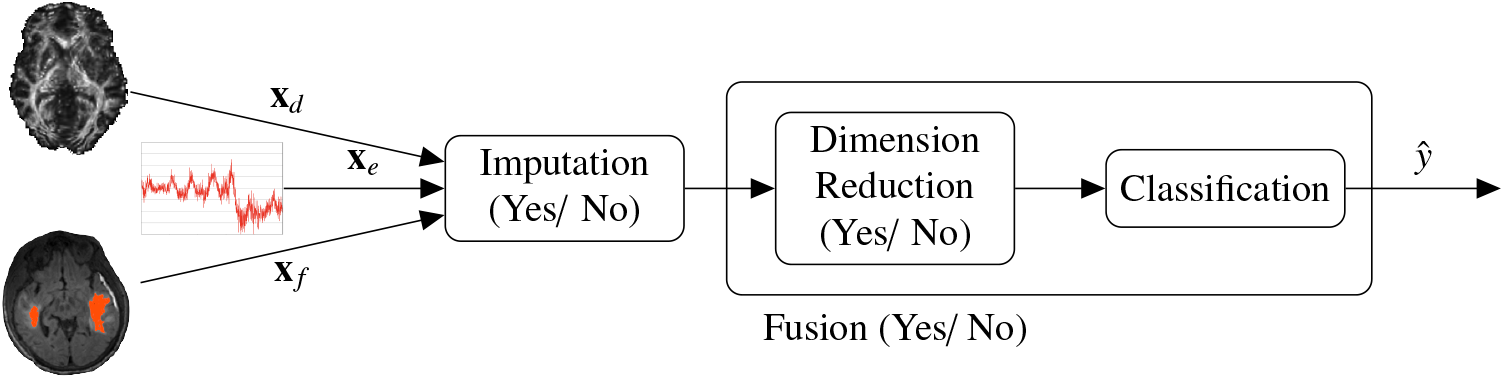
The overall modeling framework for the seizure classification task. High-dimensional features from the three different input modalities (x_*d*_, x_*e*_, x _*f*_) are first concatenated, and then passed on to the first stage. The data is processed in each stage, and proceeds along in the direction of the arrows. The final output on the right is the predicted binary seizure label (ŷ). The *(Yes*/ *No)* choices dictate whether a certain stage is active or not: if yes, it uses one of the several techniques discussed later; if no, it becomes transparent where the output is the same as the input.

#### 2.2.3 Imputation Techniques

Most classification techniques documented in the literature are not equipped to handle missing data. To establish baselines for individual modalities and apply several relevant classification pipelines, we explored four standard techniques for imputing missing modality features: mean, median, k-nearest neighbors (kNN), and iterative imputation.

The mean and median imputation strategies substitute each missing feature in 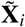 with the mean and median, respectively, of the non-missing values in the corresponding feature column of **X**_train_. For kNN imputation, each missing feature in 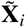 is replaced with the mean value of the feature found in a specified number of nearest neighbors *s*_ne_ (based on Euclidean distance among the non-missing features) of 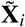 within **X**_train_ (Troyanskaya et al., 2001). Iterative imputation, on the other hand, fills the missing values in 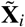 by modeling the *d* features with missing values as a function of the other *D−d* features in a roundrobin fashion, for a predefined number of iterations. For all the imputation techniques utilized, we extracted information solely from **X**_train_ (in an unsupervised learning environment), and not from **y**_train_.

#### 2.2.4 Dimensionality Reduction and Classifiers

When considering **X**_train_ as input, the feature dimension *D* is considerably larger than *n*. Aside, by the inclusion of all features, we risk hurting the final classification performance by adding noise and overfitting by increasing complexity (Kohavi and John, 1997). Consequently, it is advisable to reduce the dimensionality of such high-dimensional input. As such, we cascaded a standard dimension reduction technique at the start of each classifier for all fusion approaches, except for the intermediate fusion techniques (Other) outlined in Sections 2.6.4-2.6.7, which inherently serve the purpose of dimension reduction. The three standard reduction procedures adopted in this work are: principal component analysis (PCA) (Hotelling, 1992), and *K*best features chosen by maximizing either the χ^2^-test score 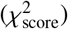 or the Fischer score (*f*_score_), between **X**_train_ and **y**_train_ (Pe-dregosa et al., 2011).

For all classification tasks, either as an interim individual learner or as the final estimator, we evaluated both SVM and AdaBoost classifiers. The choice of these two classifiers aligns with previous studies, which have found both SVM (Jie et al., 2015) and AdaBoost (Akbar et al., 2021) to be suitable for classification tasks involving neuroimaging features. For SVM, we explored both linear and radial basis function (RBF) kernel variants.

The top *K* or *k* features/components to be chosen, for either one of the standard techniques or the intermediate fusion techniques, were varied in the search space as shown in Fig. 2. The implementation difference lies in the selection of different values for *K* per CV fold for the standard reduction techniques, following a nested 5-fold CV within the training set, whereas the same value for *k* was utilized (for computational considerations) for all outer CV folds involving the intermediate reduction techniques. For the classifiers, the search space for SVM RBF’s gamma (γ), L-2 regularization (𝓁_2_), and the number of trees (*tree*) in AdaBoost can also be seen in Fig. 2. All models were trained and tested in Python 3.7.4, with support from scikit-learn (Pedregosa et al., 2011) version 1.0.2, CCA-Zoo(Chapman and Wang, 2021), and MMIDimReduction (Özdenizci and Erdoğmuş, 2021). The source code for this work is open-source and available to the public. ^1^

**Fig. 2.**
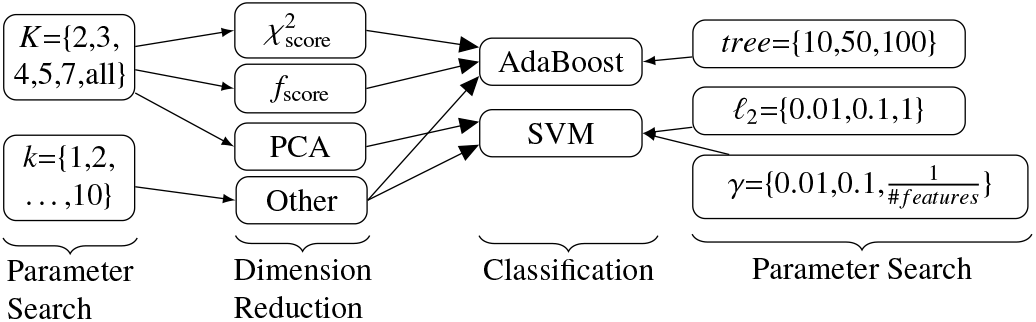
Model pipeline showing the different dimension reduction techniques and the classifiers, along with their parameter search spaces. The *Other* block within dimension reduction contains our extension of an existing technique (Section 2.3), as well as our two proposed algorithms (Sections 2.6.6 and 2.6.7).

### 2.3 No Imputation: Extended Naive Bayes (NB-extend) Late Fusion Model

In this section, we propose an extension to the conventional Naive Bayes (NB) classifier (Murphy, 2012) within our graphical model framework to address missing modalities in the original, incomplete dataset. As illustrated in Fig. 3, we postulate that the resulting physical state of the subject is captured by the different observed modalities (*x*_*d*_, *x*_*e*_, *x* _*f*_), given the LPTS label (*y*). The intent of multimodal fusion using the different modalities is to maximize the likelihood of accurately identifying the true LPTS label, based on the evidence gathered. In our NB-extend approach, we consider the features from each modality to be conditionally independent of one another, such that

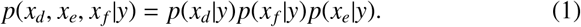

Under the conditional independence assumption, it is possible to integrate out the contribution of any missing modalities, i.e., by marginalizing over all possible values of *x* _*j*_ given *y*, where *j* ∈ *M* and *M* = { *d, e, f* } . This formulation can be directly extended to the missing modality scenario. For instance, suppose *x*_*e*_ is the missing modality, Eq. (1) becomes *p*(*x*_*d*_, *x* _*f*_ | *y*) which is equivalent of marginalizing over all possible values of *x*_*e*_ given *y*, leading to:

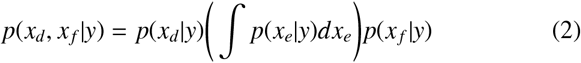

where ∫ *p*(*x*_*e*_ | *y*)*dx*_*e*_ = 1.

**Fig. 3.**
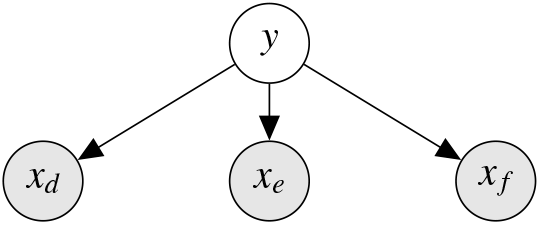
Graphical model depicting the hypothesized relation of the observed modalities (shaded circles: *x*_*d*_, *x*_*e*_, *x* _*f*_), with LPTS label as the latent state (clear circle: *y*).

The maximum a-posteriori estimation of choosing the predicted label ŷ of a given subject is then taken to follow

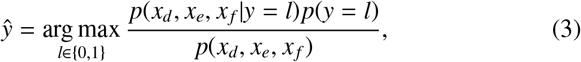

where *l* = 0 (no LPTS), *l* = 1 (LPTS), and

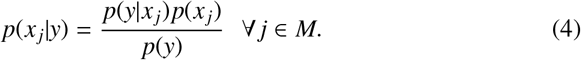

Since *p*(*x*_*j*_) and *p*(*x*_*d*_, *x*_*e*_, *x* _*f*_) are not functions of *y*, the optimization objective in (3) is equivalent to

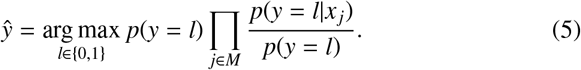

In this work, output from each individual modality base learner (elaborated in Section 2.4) was used to approximate *p*(*y*| *x*_*i*_), and *p*(*y* = *l*) is estimated from the train set.

### 2.4 With Imputation: No Fusion Model

We developed *M* individual modality base learners, using the data with imputation and without explicit fusion (disregarding the cross-modality information lookup during imputation). The imputed features from each *j*-th (*j* ∈ *M*) modality were passed through a standard reduction technique and then fitted to a classifier, as outlined earlier in Fig. 2.

### 2.5 With Imputation: Late Fusion Model

In this section, we will discuss the implementation of the late fusion ensemble classification on the imputed data. We begin by collecting the probabilities *p*(ŷ _*j*_) corresponding to the predictions

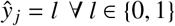

from each *j*-th modality classifier (base learner), as introduced in Section 2.4. We then summed these probabilities for *l* = 0 and *l* = 1, and utilized maximum likelihood to estimate

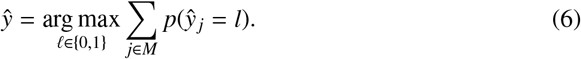

This manner of probabilistic fusion (soft) takes into account the confidence in the predictions of the individual base learners. This is in contrast to another comparable implementation, where each learner directly estimates the output label, and the fusion ultimately involves only majority voting (hard) among the estimated labels (Wolpert, 1992).

### 2.6 With Imputation: Intermediate Fusion Models

In this section, we will discuss the implementations of several existing intermediate fusion techniques, as well as propose two novel techniques at the end.

#### 2.6.1 Feature Union

In this approach, all *D* features from the three modalities were concatenated to give **X**_train_ ∈ ℝ^*n*×*D*^. Subsequently, **X**_train_ was passed through a standard reduction technique to extract *K* features, and then fitted to a classifier.

#### 2.6.2 Sequential Feature Selection (SFS)

For the selection of features from such high *D* dimensional data, standard univariate feature selection techniques ignore the mutual information among features. Multivariate feature selection techniques, however, consider a group of features in its entirety. Unfortunately, searching for the globally optimal subset with exhaustive search is *O*(2^*n*^), and can be computationally intractable. As a result, it is common practice to resort to algorithms striving to obtain a locally optimal, but perhaps globally sub-optimal, feature set with a lower complexity (Dash and Liu, 1997). The wrapper method, a multivariate feature selection technique, attempts to find a set of highly important features by fitting a particular classifier on all features in a nested CV within the train set (Erdogmus et al., 2007). Wrapper methods have been demonstrated to provide superior performance (compared to filter methods in feature selection) in classification tasks involving high-dimensional neuroimaging data with limited samples (Jie et al., 2015).

In SFS, a multivariate wrapper technique, we greedily choose features for classification, either by forward selection or by backward elimination. For the first round in forward selection, based on the score of the wrapper classifier, the most informative feature is added. In the case of backward selection, the least informative feature is removed. Forward selection thus involves a bottom-up search strategy, which begins with an empty set, and during each iteration, a new feature is added to the current set so the loss function is reduced (Kohavi and John, 1997). On the contrary, backward elimination, follows a top-down approach, starting with the complete set *D*, features are removed one at a time such that the reduction in performance is kept at a minimum (Kohavi and John, 1997). Choosing either forward or backward selection, we retain *k*_SFS_ features. Selected features are collected as 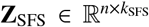, and subsequently fitted to a final classifier. SFS has an average complexity of O(*n*^2^).

#### 2.6.3 Stochastic Mutual Information Gradient (SMIG)

In this method, we aim to learn a feature transformation network 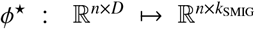 such that the high *n* × *D*-dimensional input feature space is mapped to a lower *n* × *k*_SMIG_-dimensional transformed feature space (Özdenizci and Erdoğmuş, 2021). This mapping is done while maximizing the mutual information between the transformed set and the train labels **y**_train_ as

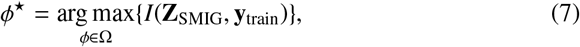

where *I*(·) is the information/ mutual information function, **Z**_SMIG_ contains the transformed training samples, and Ω denotes the function space for possible feature mappings ϕ. The parameters of this ϕ^⋆^(·) network is updated iteratively by a technique known as SMIG (Özdenizci and Erdoğmuş, 2021).

#### 2.6.4 Canonical Correlation Analysis (CCA)

We hypothesized that by looking in the directions of the maximal correlations of the input modalities, we would be able to extract information about their shared space, which might help the classification task, since the *y* was assumed to be the underlying common latent state in Fig. 3. In CCA, two sets of variables are linearly combined to obtain canonical variates such that the correlation between the linear combinations is maximized (Hotelling, 1992). This technique was thus utilized to combine our input modalities in a three-fold combination: (*x*_*d*_, *x*_*e*_); (*x*_*d*_, *x* _*f*_); and (*x*_*e*_, *x* _*f*_).

For mathematical formulation, let us assume the combination (*x*_*d*_, *x* _*f*_). If we can express two linear functions *u* = *a*^T^*x* _*f*_ and *v* = *b*^T^*x*_*d*_, and if the covariance (Hotelling, 1992)

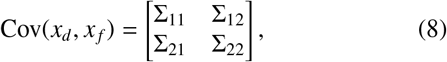

it can be shown that their correlation is maximized when

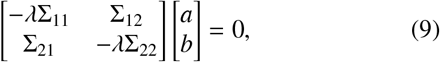

where the eigenvalues λ ∈ { ρ_1_≥ … ≥ ρ_*s*+*t*_ } are in descending order. The maximum correlation was obtained with λ = ρ_1_, and it provided our first pair of variates 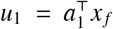 and 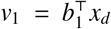. From the obtained *D* such pair of variates, we then selected *k*_CCA_ pairs to form 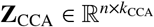, and fitted **Z**_CCA_ to a final classifier.

#### 2.6.5 Generalized Canonical Correlation Analysis (GCCA)

CCA can only linearly combine two sets of variables. GCCA, however, can combine more than two variables (Tenenhaus and Tenenhaus, 2011). In our case, GCCA combined (*x*_*d*_, *x*_*e*_, *x* _*f*_) directly. GCCA attempts to solve the following optimization problem

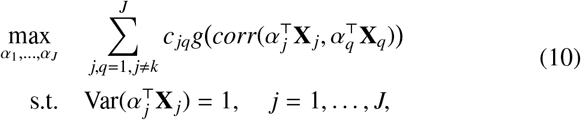

where *corr*(·) is the Pearson correlation function, *g*(·) is the identity function, *c* _*jq*_ = 1 represents the connections between modality feature matrices { *x*_*d*_, **X**_*e*_, **X** _*f*_ }, and *J*=3.

Similar to CCA, we then selected *k*_GCCA_ pairs of variates from the total *D* pairs, to give 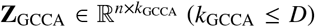, and fitted **Z**_GCCA_ to a final classifier.

##### Algorithm 1: RECC for a given selection algorithm

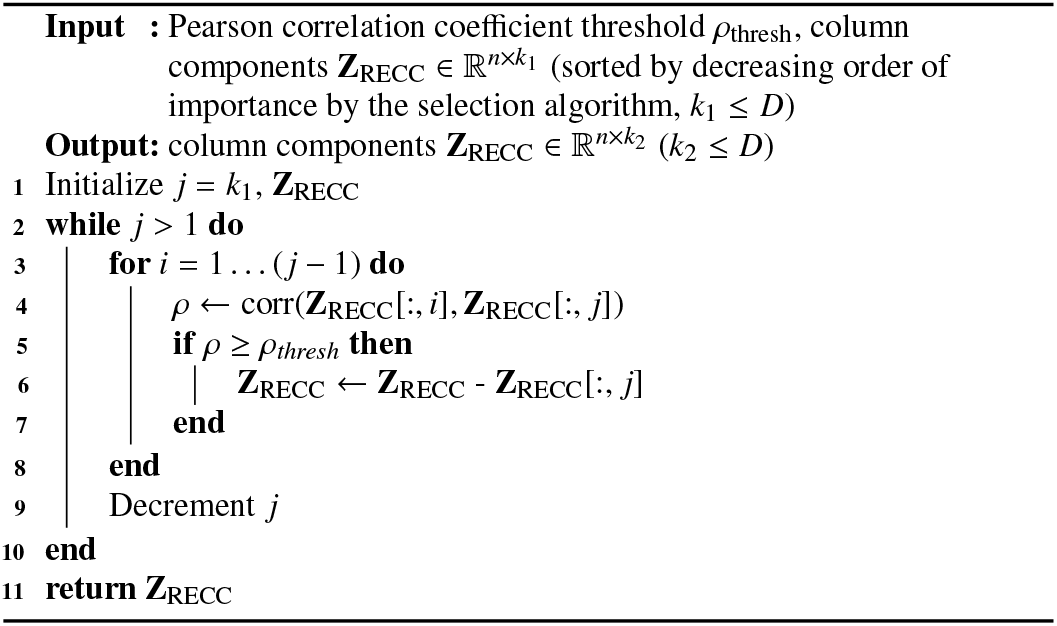

#### 2.6.6 Recursive Elimination of Correlated Components (RECC)

##### Background

In CCA, each of the *k*_CCA_ pairs of canonical variates chosen is uncorrelated to each other, since the pairs maintain orthogonality among themselves (Hotelling, 1992). However, there is no such imposed constraint on some of the other dimension reduction techniques like SFS or SMIG. As such, there exists a possibility that the chosen set of *k* features/projections (components) will likely contain a set of components, which are highly correlated with each other. The inclusion of such components might negatively impact the classification performance (Kohavi and John, 1997).

##### Proposition

To limit the inclusion of redundant components, we introduce a novel technique, RECC, designed to exclude less critical but highly correlated components from our analyses. As outlined in Algorithm 1, our approach initiates with a set of components 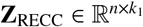, arranged in descending order of importance based on a predetermined selection algorithm. Starting with the component of least importance, we compute the Pearson correlation coefficient (ρ) of this component against all other components of higher importance. If the computed coefficient reaches or surpasses a predefined threshold (≥ ρ_thresh_), we eliminate this component from **Z**_RECC_. We continue this comparison, and elimination if applicable, until all components in **Z**_RECC_ have been analyzed. The remaining components in 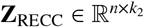 are then fitted to the final classifier.

#### 2.6.7 Information Decomposition and Selective Fusion (IDSF)

##### Background

Our earlier hypothesis in Section 2.6.4 was that by utilizing information from the shared space among the

###### Algorithm 2: IDSF for one decomposition algorithm and one selection algorithm prepared by RECC

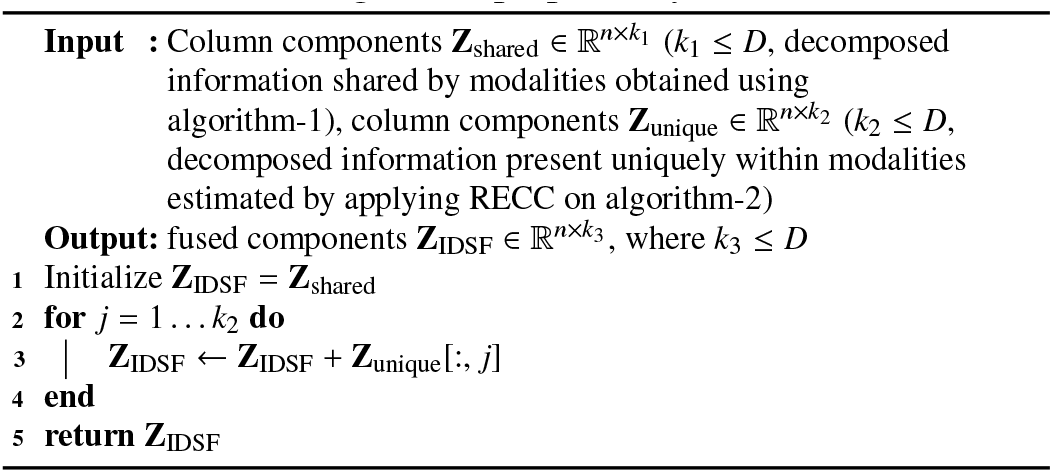

modalities, we would hope to observe a better classification performance, since the LPTS label *y* was also assumed to be a shared latent state. However, the possibility exists that a considerable fraction of the information, useful for classification, is present uniquely in a space not shared by the individual modalities (Williams and Beer, 2010). This is illustrated in Fig.4. The total information provided by two sample modalities *x*_1_ and *x*_2_ about *y* may be denoted by the mutual information measure *I*(*y*; *x*_1_, *x*_2_). However, it is also important to understand how exactly *x*_1_ and *x*_2_ contribute to this aggregate information. Three primary scenarios may arise (Williams and Beer, 2010) when examining the individual contributions of *x*_1_ and *x*_2_. Firstly, *x*_1_ and *x*_2_ may offer common shared information as

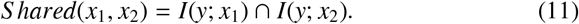

where ∩ is the intersection operator. Secondly, *x*_1_ might offer unique information, *Unique*(), not provided by *x*_2_, or vice versa. An example of the former case is

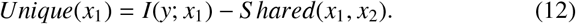

Finally, the combination of *x*_1_ and *x*_2_ may provide synergistic information that is not available from either alone, but only obtained when both are considered together (e.g. an XOR gate for binary inputs)

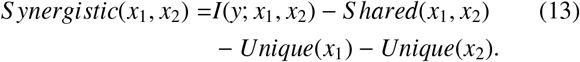

**Fig. 4.**
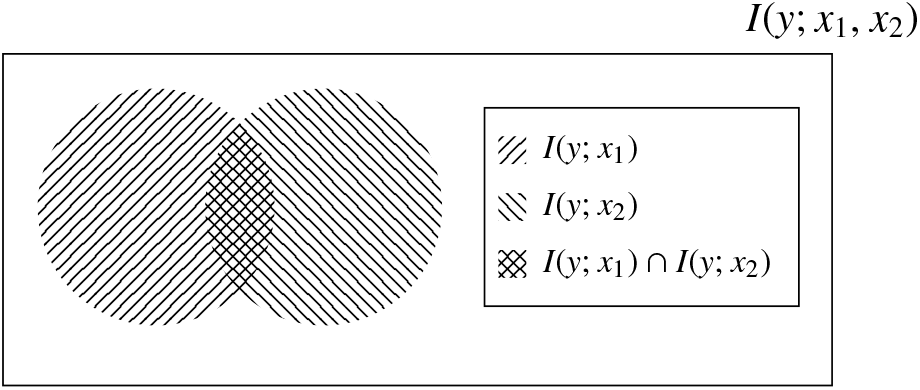
Structure of multivariate information that two sample modalities *x*_1_ and *x*_2_ would likely provide about the LPTS label *y*.

##### Proposition

The preceding discourse highlights the potential benefits a classifier may accrue from an amalgamation of shared and unique information derived from multiple modalities. We chose not to delve into synergistic information, due to the inherent complexities associated with its extraction. Building upon this multivariate information structure, we lay the groundwork for our proposed IDSF algorithm. This method aims to integrate shared information, represented by **Z**_Shared_ (for instance, canonical variates sourced from CCA), with the estimated unique information symbolized by **Z**_Unique_ (such as components singled out by RECC). This amalgamation results in **Z**_IDSF_, which is subsequently fitted to the final classifier. A detailed depiction of the IDSF technique is provided in Algorithm 2.

### 2.7 Evaluation Metrics

*Model Performance*: For evaluating models on the test partitions in each CV fold, we use mean area under the receiver operating characteristic (ROC) curve (AUC) as the primary metric. Hanley et. al. demonstrated AUC to be an effective measure of accuracy, with a meaningful interpretation in medical diagnosis (Hanley and McNeil, 1982). We thus used AUC to rank the models. Aside, AUC helps choose a desired operating point (sensitivity, specificity) in the ROC curve, where

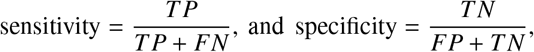

where *T P, FN, TN*, and *FP* are the true positive, false negative, true negative, and false positive, respectively. Two techniques have been suggested in literature (Hajian-Tilaki, 2013) to choose the best operating point (for equal weights to sensitivity and specificity), including the highest

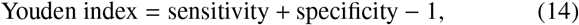

and the lowest Euclidean distance from the upper left corner (0,1) of the ROC curve, as

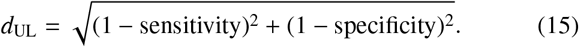

As a secondary metric to assess model performance on this slightly imbalanced dataset, but not to rank them, we used weighted F1-score as given by

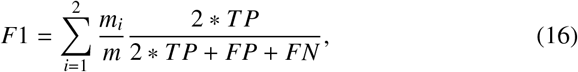

where *m*_*i*_ is the number of samples belonging to the *i*-th class, in the current test fold.

#### Feature Contribution

For interpreting the top models, the SHapley Additive exPlanations (SHAP) framework is used. Introduced by (Lundberg and Lee, 2017), it builds upon the concept of Shapley values from cooperative game theory. By using SHAP values, one can fairly distribute the contribution of each feature to a prediction for a specific instance. These values enable a better understanding of the inner workings of complex ML models, such as decision trees, SVMs, and even deep learning networks. Mathematically, the SHAP value ϕ _*j*_ of feature *j* is computed as the average marginal contribution of the feature across all possible combinations of features:

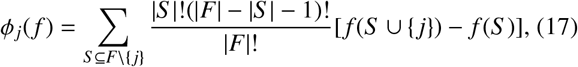

where *F* is the set of all features, *S* indicates all feature subsets obtained from *F* that do not include feature *j*, and *f* is the ML model. We use the kernel method of SHAP calculation (Lundberg and Lee, 2017).

## 3. Results

### 3.1 Model Performance

#### Imputation

Based on our preliminary classification results with the four different imputation techniques, kNN imputation outperformed the other three. For this reason, all subsequent results in this section, wherever data was imputed, was done using the kNN imputer, with *s*_ne_ = 1.

#### NB-extend

The comparison of the mean AUC and the mean weighted F1 performances, on the test sets from all five CV folds, for the different fusion techniques (except IDSF) are summarized in Table 2. The first row shows the classification performance of our extended NB-extend estimator with late fusion, where the original input features (no imputation) were used to fit individual modality base learners. NB-extend (AUC=0.710) outperformed the individual modality base learners (AUC= {0.664,0.670,0.619}), using imputed features without any fusion, and which form the next three rows of Table 2.

#### Late Fusion

Of the techniques using imputed features and fusion, we first list the performance of the soft late fusion estimator. This probabilistic estimator also performed better (AUC=0.756) than the individual modality base learners. It even performed better than both the late fusion NB-extend estimator (no imputation), and the intermediate fusion (union of all modality features) classifier (AUC=0.736), in the subsequent row of Table 2.

#### Intermediate Fusion

The last six rows of Table 2 correspond to the dimension reduction techniques involving intermediate fusion, as discussed earlier in Sections 2.6.4-2.6.6. Among them, the first four rows presented existing techniques in literature, and the best performance (AUC=0.784) was achieved with CCA. Even though GCCA took into account all three modalities (*x*_*d*_, *x*_*e*_, *x* _*f* −*n*_), compared to CCA’s two (*x*_*d*_, *x* _*f* −*p*_), we noticed it lagged in performance (AUC=0.654) substantially compared to that of CCA. Of the two SFS implementations discussed (forward and backward), forward selection performed better (AUC=0.676), and was thus recorded. Similarly, between the two implementations of SMIG (linear and non-linear), linear performed better (AUC=0.699) and was recorded. Finally, the last two rows in Table 2 demonstrated the improvement in performances with our proposed RECC (AUC= { 0.710, 0.753}), when applied on the original SFS and SMIG implementations, respectively. The best F1 score (0.753) was obtained with RECC-SMIG, indicating a potential choice for the best model, if F1 score is more important in the model selection.

#### Proposed RECC

To obtain a better understanding of how the choice of ρ_thresh_ affected the AUC performances of the RECC combinations corresponding to the last two rows in Table 2, we plotted AUC vs. ρ_thresh_ in Fig. 5. The standard implementations of SFS and SMIG corresponded to ρ_thresh_ = 1. As we lowered ρ_thresh_, we noticed the AUC performances go up until they peaked at ρ_thresh_ = 0.5 for both RECC implementations, and then started to go down. At ρ_thresh_ = 0.15, the performance of RECC-SMIG almost returned to its SMIG baseline, whereas that of RECC-SFS remained higher than its SFS baseline.

**Table 2.**
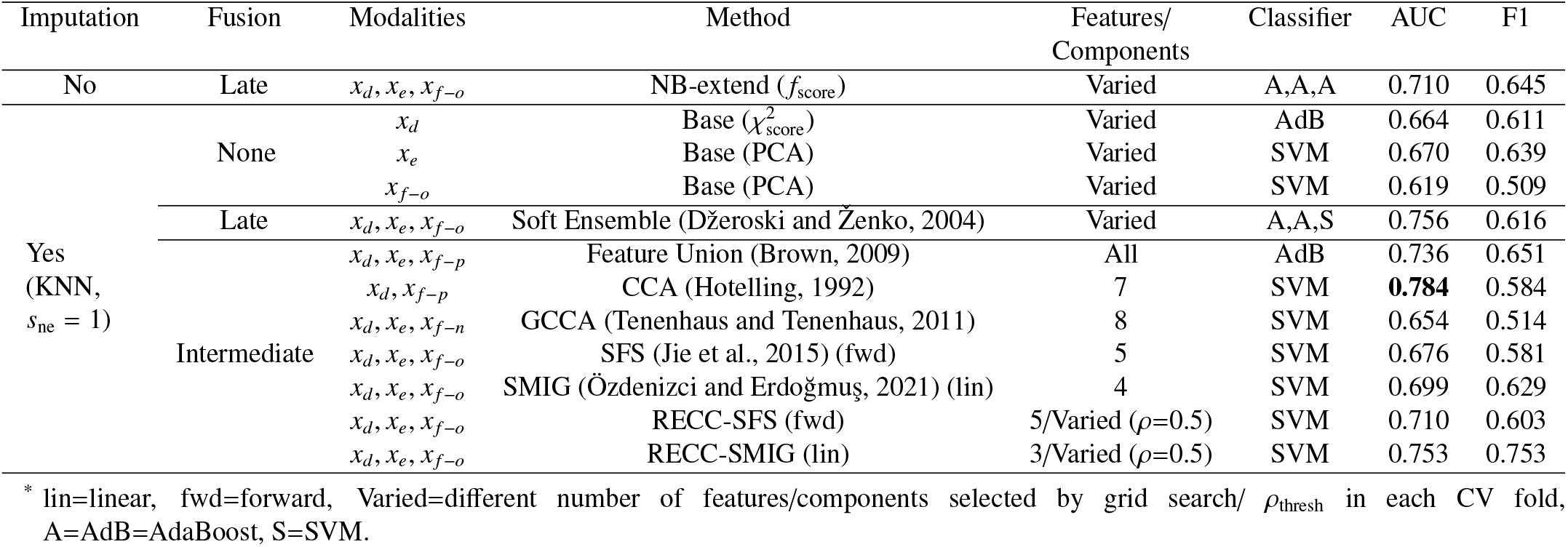
Performance of the different dimensionality reduction techniques, for different stages of fusion, with both original and imputed features.

**Fig. 5.**
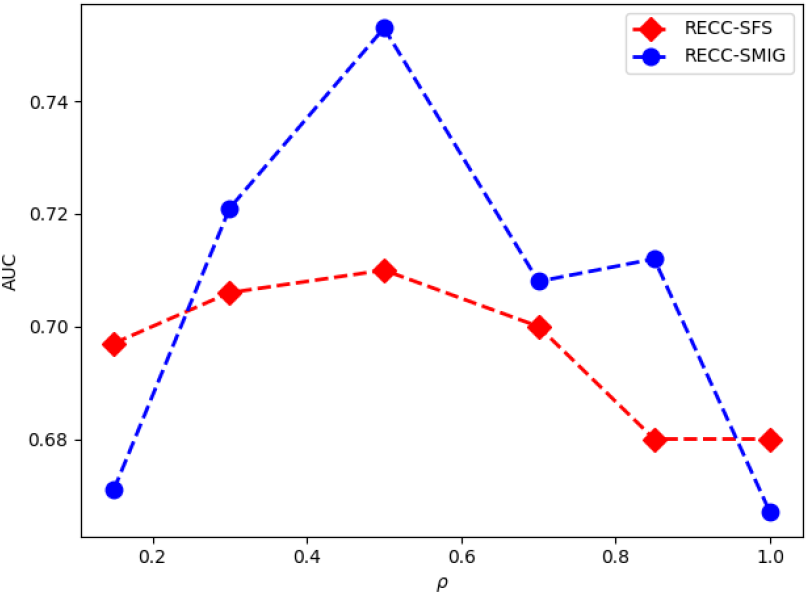
AUC performance of RECC-SFS and RECC-SMIG combinations from the last two rows in Table 2, as a variation of the Pearson coefficient threshold ρ_thresh_, with SVM classifier and modalities *x*_*d*_, *x*_*e*_, *x* _*f* −*o*_.

#### Proposed IDSF

In Table 3, we recorded the best AUC performances for each combination of the two decomposition algorithms (CCA and GCCA) with the two selection algorithms (SFS and SMIG) prepared by RECC, using intermediate fusion with IDSF. We found the best IDSF performance (AUC=0.792), also the best among all tested models, was obtained with CCA and RECC-SFS. Just as seen earlier with CCA and GCCA, IDSF with CCA was superior to IDSF with GCCA. Interestingly, while individually both SMIG and RECC-SMIG performed considerably better than SFS and RECC-SFS earlier, their IDSF performances were closely matched. Aside, while IDSF improved on the performances of the decomposition algorithms on all occasions, it did not necessarily do so over that of the selection algorithms (e.g. IDSF with GCCA and RECC-SMIG). Finally, the best F1 score (0.677) for IDSF was obtained with GCCA and RECC-SFS.

**Table 3.** Performance of IDSF (with its inputs) in intermediate fusion with imputed features.

| Modalities | Shared | Components | Modalities | Unique | Components | Classifier | AUC | F1 |
| --- | --- | --- | --- | --- | --- | --- | --- | --- |
| $x_d, x_{f-p}$ | CCA | 7 | $x_d, x_e, x_{f-o}$ | RECC-SFS (fwd) | 5 (Varied, $\rho=0.5$) | SVM | **0.792** | 0.619 |
| $x_d, x_{f-p}$ | CCA | 6 | $x_d, x_e, x_{f-o}$ | RECC-SMIG (lin) | 1 (Varied, $\rho=0.5$) | SVM | 0.783 | 0.584 |
| $x_d, x_{f-p}$ | GCCA | 1 | $x_d, x_e, x_{f-o}$ | RECC-SFS (fwd) | 5 (Varied, $\rho=0.5$) | AdB | 0.663 | 0.677 |
| $x_d, x_{f-n}$ | GCCA | 8 | $x_d, x_e, x_{f-o}$ | RECC-SMIG (lin) | 4 (Varied, $\rho=0.5$) | AdB | 0.670 | 0.642 |

\* lin=linear, fwd=forward, Varied=different number of components were selected by $\rho_{\text{thresh}}$ in each CV fold, AdB=AdaBoost.

#### ROC Analysis

To examine the effect of the proposed algorithms RECC and IDSF in terms of operating points, mean ROC curves (from the 5-fold CV) are plotted in Fig. 6 for the best model from Table 3, as well as for the individual techniques that went into the best model from Table 2. The variation of the best ROC curve (IDSF) is estimated by a binomial distribution approximation, for a 95% level of confidence (Sourati et al., 2015). We noticed that just like RECC-SFS improved on standard SFS, IDSF (AUC=0.79) improved on the performance over its two input techniques: CCA (AUC=0.78) and RECC-SFS (AUC=0.71). It also obtained the best operating point (0.01, 0.67: shown in a red circle), according to the highest Youden index (0.68) and the lowest *d*_UL_ (0.23).

### 3.2 Feature Importance

*Wilcoxon Rank-Sum Test*: In a bid to evaluate if the most impactful features from the classification tasks could also serve as potential biomarkers, we compared the distribution of these frequently selected features across the two LPTS groups. These features, derived from the top-performing algorithm on each dataset (both original and imputed), were aggregated from all five cross-validation folds. Table 4 showcases this comparison through a two-tailed non-parametric Wilcoxon rank-sum test. The most frequent features from various modalities, along with their Bonferroni corrected *p*-values (Dunn, 1961), are listed in Table 4. Notably, both cluster 90 (right middle temporal gyrus) in *x* _*f* −*o*_, chosen by NB-extend using original features, and SS–R (sagittal stratum right) in *x*_*d*_, chosen by IDSF from imputed features, achieved statistical significance at the 5% level (*p*<0.05). However, post-adjustment for multiple comparisons through the Bonferroni correction, none of the shortlisted features exhibited significant differences between the two seizure groups.

**Table 4.** Comparison of selected features between the two LPTS groups using a two-tailed Wilcoxon rank-sum test.

| Imputation | Subjects | Model | Modality | Potential Biomarker(s) | Unadjusted $p$-value | Comparisons | Corrected $p$-value |
| --- | --- | --- | --- | --- | --- | --- | --- |
| No | 41 | NB-extend | $x_d$ | ALIC-L,FX/ST-R,IFO-R | >0.05 | 3 | >0.05 |
| | 32 | NB-extend | $x_{f-o}$ | Clusters (37,48,57,66) | >0.05 | 5 | >0.05 |
| | 32 | NB-extend | $x_{f-o}$ | Cluster 90 | 0.025 | 5 | 0.125 |
| Yes | 48 | IDSF (CCA-RECC-SFS) | $x_d$ | SS-R | 0.047 | 2 | 0.094 |
| | 48 | IDSF (CCA-RECC-SFS) | $x_{f-o}$ | Cluster 48 | >0.05 | 2 | >0.05 |

\* The $p$ -values were first calculated for the two seizure groups for each feature at a 5% level of significance ( $p < 0.05$ ), and subsequently adjusted using Bonferroni correction.
Regions ( $x_d$ ): ALIC-L = anterior limb of internal capsule left, FX/ST-R = fornix/stria terminalis right, IFO-R = inferior fronto-occipital fasciculus right, and SS-R = sagittal stratum (including inferior longitudinal fasciculus and inferior fronto-occipital fasciculus) right.
Clusters ( $x_{f-o}$ ): 37 = left middle cingulate and paracingulate gyri, 48 = right calcarine fissure and surrounding cortex, 57 = left inferior occipital gyrus, 66 = right inferior parietal gyrus (excluding supramarginal), and 90 = right middle temporal gyrus.

#### SHAP Analysis

To delve deeper into the role of individual features in LPTS prediction, we employed the SHAP toolbox (Lundberg and Lee, 2017), fitted with the **X**train from each fold to interpret the **X**test for the corresponding fold. The leading models without and with imputation are NB-extend and IDSF (CCA-RECC-SFS), as earlier seen in Table 4. It is worth noting that SHAP values cannot be computed for missing entries in NB-extend, and IDSF only offers access to a limited number of features per fold. To establish a middle ground for easy comparison between these models, SHAP values were computed on the imputed data (used by IDSF), utilizing the AdaBoost classifier (same configuration as NB-extend). The resultant SHAP values for all the five folds (the entire dataset with *x*_*d*_, *x*_*e*_, *x* _*f* −*o*_) are depicted in Fig.7. The mean SHAP values illustrated in Fig.7(a) represent the overall influence of individual features, whereas the swarm plot in Fig.7(b) demonstrates the effects of the value of individual feature instances on predicting the binary LPTS labels. From Fig.7(a), it is observed that ALIC-L (anterior limb of internal capsule left) from *x*_*d*_ had the most profound impact and was also previously identified as a potential biomarker in Table 4. Fig.7(b) further reveals that higher values of ALIC-L tended to predict no LPTS, while lower values predicted LPTS. Cluster 90 from *x* _*f* −*o*_ had the second most significant effect on the seizure outcome prediction task, as it previously possessed the smallest unadjusted *p*-value among all tested features in Table4. Furthermore, Fig. 7(b) indicates that higher values of cluster 90 suggest potential LPTS, while lower values suggest otherwise.

### 3.3. Biomarker Visualization

Given the significant influence and clear implications of ALIC-L on model prediction as depicted in Fig.7, we selected this potential biomarker for visualization. Fig.8 portrays the dMRI FA maps of two subjects within the MNI space: Subject 32 exhibits one of the highest values of ALIC-L (0.64, indicating no LPTS), while Subject 4 shows one of the lowest (0.41, indicating LPTS). White contours delineate the boundaries of ALIC-L, and yellowish-green contours indicate the lesion locations. High values of ALIC-L, corresponding to a brighter tract and signifying robust white matter connectivity in Subject 32, suggest the absence of LPTS. Conversely, low values of ALICL, corresponding to a darker tract and signifying compromised white matter connectivity in Subject 4, denote LPTS. Interestingly, even though the traumatic lesion is closer to ALIC-L in Subject 32 compared to Subject 4, this proximity does not seem to affect the value/ brightness of ALIC-L, hence, not impacting its connectivity.

## 4. Discussion

### 4.1 Clinical Diagnosis

In Figure 6, we note superior AUC performance from our proposed IDSF algorithm, albeit the marginal improvement over CCA (with an AUC of 0.79 compared to 0.78). However, the operating points of the two curves vary substantially, as depicted in Figure 6. The choice of the optimal operating point often relies on clinical decision-making factors. These can range from the importance of correctly diagnosing LPTS (prioritizing sensitivity), correctly identifying no LPTS cases (prioritizing specificity), or maintaining an equivalent cost-benefit ratio for accurate and erroneous diagnoses (equating sensitivity and specificity) (Hajian-Tilaki, 2013).

**Fig. 6.**
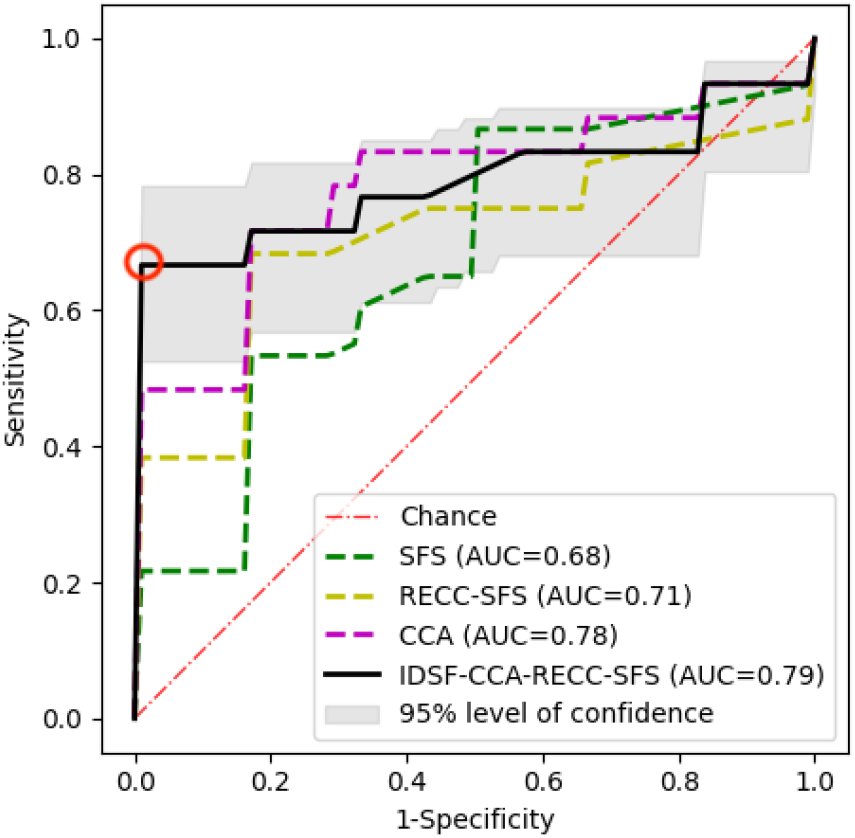
ROC curve for the best performing model (solid black line) using IDSF-CCA-RECC-SFS. The ROC plots for the classification performance with SFS, RECC-SFS, and CCA are also shown: as purple, yellow, and green dotted lines, respectively. The best operating point (as per highest Youden index=0.68 and lowest *d*_UL_=0.23) is shown in a red circle. The classifier is SVM and information from all three modalities is used.

In scenarios where the latter two cases hold greater significance, the IDSF could be considered the superior model, given its operational point demonstrated in the red circle in Figure 6. This point corresponds to the highest Youden index and the minimal *d*_UL_, indicating the optimal balance between sensitivity and specificity. Nevertheless, determining the optimal model may necessitate a more nuanced evaluation if the cost of falsenegative diagnoses is higher.

### 4.2 Biomarker Investigation

Identifying potential biomarkers through multiple datadriven analyses has its inherent challenges. Notably, there is a risk of overstated performance results due to inappropriate evaluation frameworks (Barla et al., 2008), and a lack of robustness in identified biomarkers (Feng et al., 2004). Common approaches to tackle such concerns include employing the AUC for evaluation, implementing cross-validation, and using different sets of subjects (e.g., subjects from multiple centers) (McDermott et al., 2013): all of which we have adopted in this work.

From our experiments, dMRI alterations in ALIC-L (anterior limb of internal capsule left) had the most profound effect on the classification task in Fig. 7(a), and was also earlier captured in Table 4 as one of the most frequently selected features. Lower values of ALIC-L, indicative of a darker tract and compromised white matter connectivity, were associated with LPTS (e.g. subject 04 in Fig. 8). Abnormalities of dMRI in ALIC-R (ALIC right) have been documented earlier in subjects suffering from LPTS (Akbar et al., 2021), and that in ALIC (both left and right) in temporal lobe epilepsy (TLE) (Meng et al., 2010). ALIC, or more precisely ALIC left, is thus our strongest candidate for a potential biomarker. Aside, fMRI alterations in cluster 90 (right middle temporal gyrus) had the second most profound effect on the classification task in Fig. 7(a), while having the smallest unadjusted *p*-value in Table 4. The right temporal lobe (containing the superior, middle, and inferior temporal gyrus) has been previously reported as a brain biomarker for early PTS (EPTS) and LPTS, following shape analysis with structural MRI (Lutkenhoff et al., 2020) and EEG and structural MRI (Irimia et al., 2017).

**Fig. 7.**
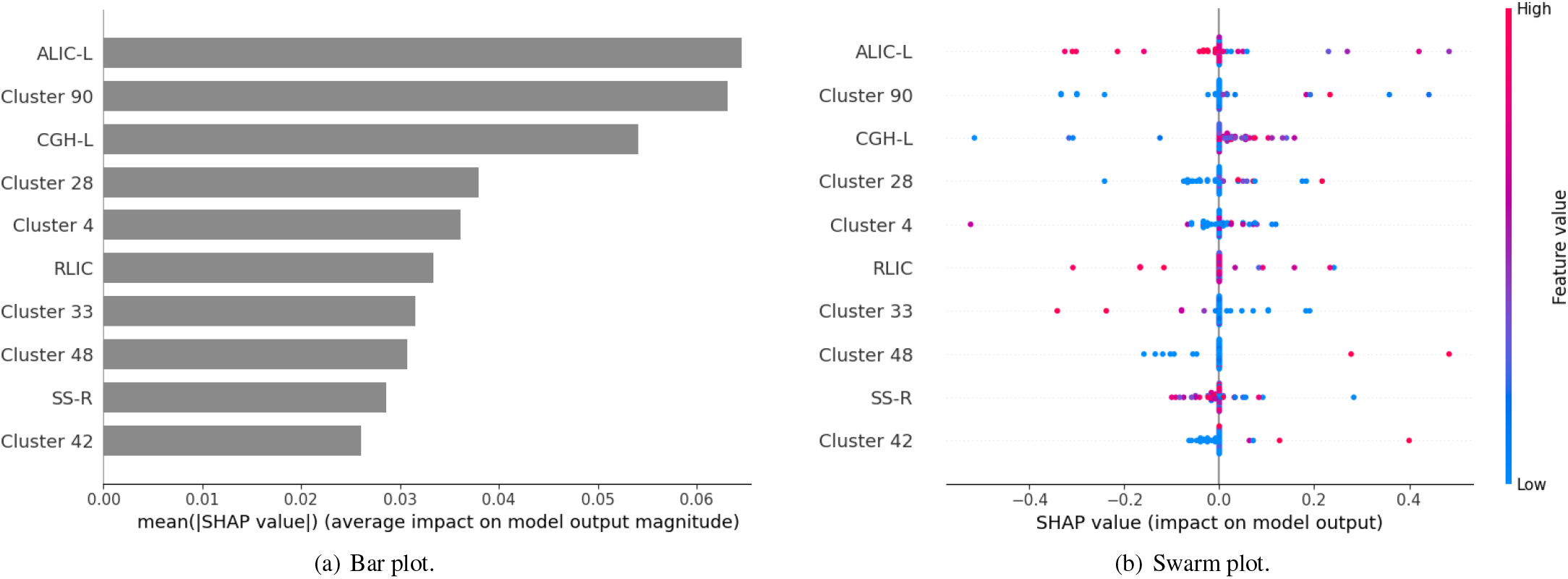
SHAP values of feature importance on the imputed data, using AdaBoost on the test sets (X_test_) from all five folds. The mean SHAP values in (a) represent the overall absolute impact of individual features, whereas (b) demonstrate the effects of the value of individual feature instances on the predictive power of the model. In (b), -0.5 imply the strongest possible impact of a feature for a prediction of no LPTS, whereas +0.5 imply the strongest possible impact for a prediction of LPTS, with 0 representing the baseline (no impact). New regions (*x*_*d*_): CGH-L = cingulum (hippocampus) left, RLIC = retrolenticular part of the internal capsule, and SS-R (sagittal stratum right). New clusters (*x* _*f* −*o*_): 4 = Superior frontal gyrus right, 28 = anterior orbital gyrus right, 33 = insula left, and 42 = hippocampus.

Interestingly, alterations in other regions, previously associated with seizure-related abnormalities, were also observed in our study, as captured in Table 4 and Fig. 7(a). For example: SS (sagittal stratum) from EEG and structural MRI has been associated with PTS (Irimia et al., 2017; Sharma et al., 2021); FX/ST (fornix/stria terminalis) from dMRI has been associated with LPTS in humans (Akbar et al., 2021) and rats (Bao et al., 2011); and CGH (cingulum hippocampus) from dMRI has been associated with LPTS (Akbar et al., 2020) and TLE (Owen et al., 2021).

**Fig. 8.**
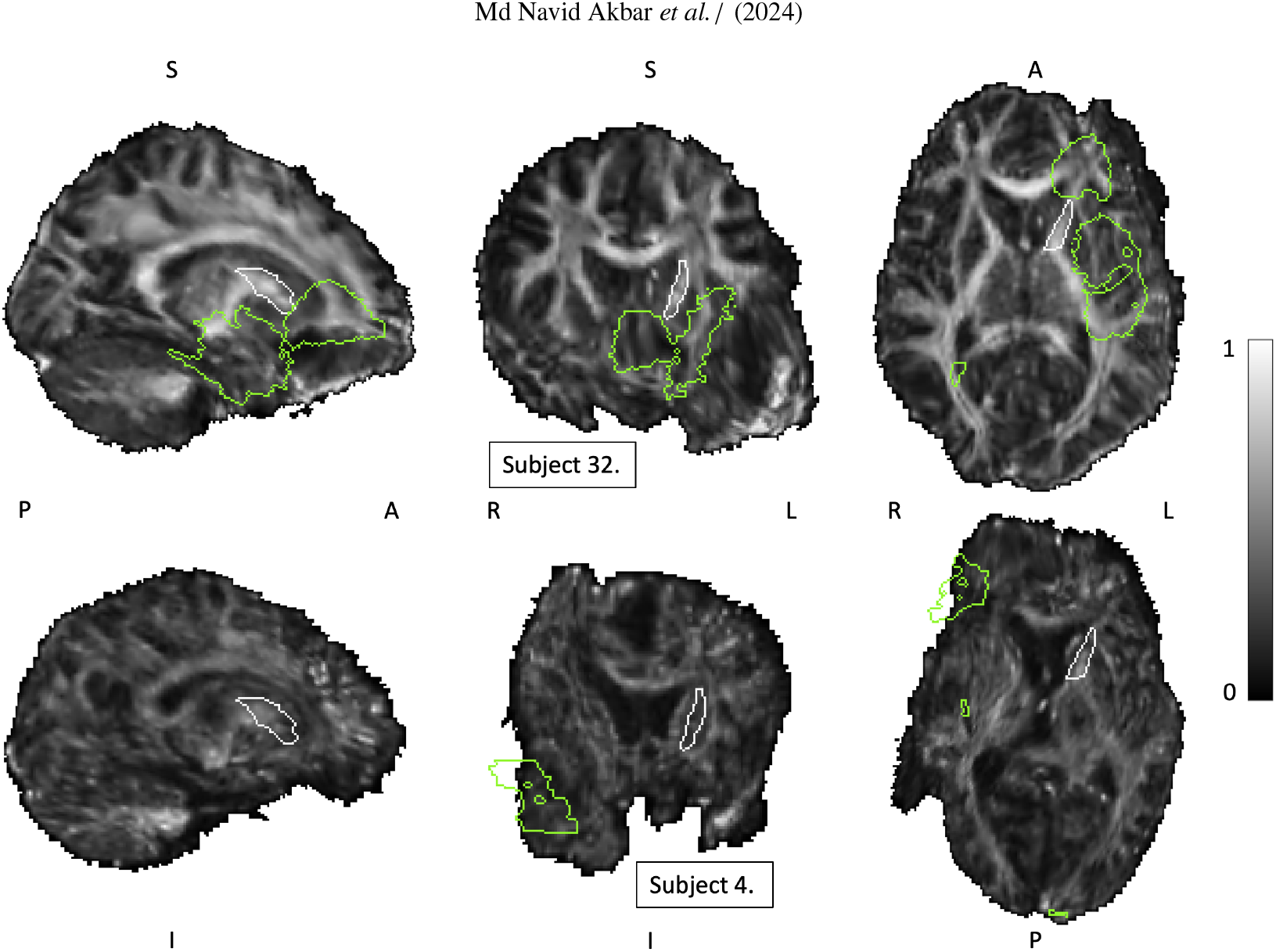
dMRI FA maps of two subjects in the MNI space: subject 32 had one of the highest values of ALIC-L (0.64, no LPTS), whereas subject 4 had one of the lowest (0.41, LPTS positive). White contours indicate the perimeters of ALIC-L, whereas yellowish-green contours indicate the perimeters of the lesion locations. The radiological directions are indicated as: P=posterior, A=anterior, S=superior, I=inferior, L=left, and R=right. The color map corresponding to all the images is shown on the right.

Our findings align with several previous studies that reported altered functional and structural connectivity in subjects with epilepsy. A direct comparison, however, is not feasible due to two key factors (La Rocca et al., 2023). First, ours is the first multimodal study investigating potential biomarkers in patients with seizures after TBI. Second, while most prior literature has explored biomarkers by comparing subjects with epilepsy and normal controls, our study focuses exclusively on a cohort of TBI patients who have or have not developed LPTS. Furthermore, our study not only consolidated and validated previous findings derived from unimodal analysis or non-TBI subjects but also advanced the field by offering additional contributions.

Specifically, our work exhibited the following:

i) Greater number of samples: Our study encompassed a substantially larger cohort of subjects than previous investigations, with 48 subjects in our analysis. This sample size surpassed the participant numbers in aforementioned TBI studies, such as 39 subjects in (Sharma et al., 2021), 33 subjects in (Irimia et al., 2017), 25 rats in (Bao et al., 2011), 22 subjects in (Akbar et al., 2021), and 16 subjects in (Meng et al., 2010). Notably, our dataset only fell short in size when compared to a non-TBI study involving 94 subjects (Owen et al., 2021).
ii) Refined localization and modalities: Our research identified more precise brain regions and modalities that warrant investigation. For instance, we established a correlation between the potential association of LPTS with greater values (lesion volume overlap) of cluster 90 in fMRI.
iii) Quantifiable impact: Leveraging an interpretable ML feature analysis, we quantified the influence of specific features on model predictions. For example, our findings revealed that the CGH-L exhibited a mean normalized impact of 0.055 on model prediction, underscoring its importance in predicting LPTS. By encompassing a larger cohort, identifying specific brain regions and modalities, and quantifying the impact of features through interpretable ML analysis, our work contributes to a more comprehensive understanding of potential biomarkers in the context of LPTS.

### 4.3 Limitations

Our study, while pioneering in its approach, acknowledges certain constraints. Given the longitudinal nature of our study, our analysis is limited to the currently available subjects who have completed the necessary two-year follow-up (Temkin, 2009). As EpiBioS4Rx is projected to enroll a total of 300 subjects upon completion, we anticipate a substantial increase in our sample size. This enhanced dataset will permit robust testing of our proposed methods and provide the opportunity to investigate other cutting-edge techniques, such as EmbraceNet (Choi and Lee, 2019). Despite its potential for multimodal classification in cases of missing data, EmbraceNet requires a larger sample size to effectively train its deep network, which houses significantly more parameters than the methods outlined in our current work. Furthermore, we acknowledge that the potential heterogeneity in the distribution of acquired data from the different participating centers has not been thoroughly investigated in this study. Addressing this aspect will be a focus of our future work.

Further, the impact of data imputation on the distribution of the original features remains an unexplored facet in this study, and will be addressed in future work. Although our proposed IDSF algorithm (utilizing imputed features) showcased superior AUC performance, NB-extend (without imputation) proved more valuable in identifying potential biomarkers. To balance these divergent outcomes, we calculated SHAP values for a hybrid model configuration, which incorporates attributes from both NB-extend and IDSF, though it does not fully embody either.

With regard to potential biomarkers, it is plausible that other candidates exist that have not been identified in this study. These may have been overlooked either due to a lack of significant group-level differences or due to their non-dominant role in our SHAP analysis, possibly owing to the variations in individual anatomical features (Singh et al., 2021).

## 5. Conclusions

In this study, we meticulously collected multicenter data of TBI subjects and embarked upon the challenging task of binary late seizure classification, leveraging key modalities (fMRI, dMRI, EEG). We proactively addressed the issue of missing data by formulating the NB-extend classifier (aligned with our hypothesized graphical model), and implementing widely recognized imputation techniques. These approaches enabled the utilization of established dimensionality reduction techniques and machine learning classifiers.

In terms of AUC performance, while the NB-extend estimator outperformed imputation-based individual modality base learners, our proposed IDSF algorithm excelled within our experimental framework. IDSF was designed to capture both shared and unique information embedded within multiple input modalities. To ascertain the unique information in each modality, we introduced RECC, another novel technique that filters out selected information based on its correlation.

Lastly, through rigorous statistical and SHAP analyses of the most frequently selected features, we found evidence supporting dMRI abnormalities in the left anterior limb of the internal capsule and fMRI alterations in the right middle temporal gyrus as potential biomarkers of LPTS. These findings may ultimately facilitate the early diagnosis and prevention of PTE, profoundly transforming patient prognosis.

## Data Availability

The data analyzed in this study is subject to the following licenses/ restrictions: access to data must be requested and approved by the EpiBioS4Rx steering committee. Requests to access these datasets should be directed to.

## Data Availability

The data analyzed in this study is subject to the following licenses/restrictions: access to data must be requested and
approved by the EpiBioS4Rx steering committee. Requests to access these datasets should be directed to.

https://epibios.loni.usc.edu/

## Declaration of Generative AI and AI-assisted technologies in the writing process

During the preparation of this work, the authors used Chat-GPT in order to improve readability and language. After using this tool, the authors reviewed and edited the content as needed and take full responsibility for the content of the publication.

## Footnotes

^1^https://github.com/neu-spiral/Epileptogenesis.

^2^The code used to generate the results is made publicly available at https://github.com/neu-spiral/Epileptogenesis.

## References

Akbar, M.N., La Rocca, M., Garner, R., Duncan, D., Erdoğmuş, D., 2020. Prediction of epilepsy development in traumatic brain injury patients from diffusion weighted mri, in: Proceedings of the 13th ACM International Conference on PErvasive Technologies Related to Assistive Environments, New York, NY, USA.

Akbar, M.N., Ruf, S., La Rocca, M., Garner, R., Barisano, G., Cua, R., Vespa, P., Erdoğmuş, D., Duncan, D., 2021. Lesion normalization and supervised learning in post-traumatic seizure classification with diffusion mri, in: Computational Diffusion MRI, Springer International Publishing, Cham. pp. 133–143.

Akrami, H., Irimia, A., Cui, W., Joshi, A.A., Leahy, R.M., 2021. Prediction of posttraumatic epilepsy using machine learning, in: Medical Imaging 2021: Biomedical Applications in Molecular, Structural, and Functional Imaging, SPIE. pp. 424–430.

Andersen, S.M., Rapcsak, S.Z., Beeson, P.M., 2010. Cost Function Masking during Normalization of Brains with Focal Lesions: Still a Necessity? NeuroImage 53, 78–84. doi:10.1016/j.neuroimage.2010.06.003.

Ashburner, J., 2009. Computational anatomy with the spm software. Magnetic resonance imaging 27, 1163–1174.

Ashburner, J., Friston, K.J., 2005. Unified segmentation. NeuroImage 26, 839–851.

Bao, Y.h., Bramlett, H.M., Atkins, C.M., Truettner, J.S., Lotocki, G., Alonso, O.F., Dietrich, W.D., 2011. Post-traumatic seizures exacerbate histopathological damage after fluid-percussion brain injury. Journal of neurotrauma 28, 35–42.

Barla, A., Jurman, G., Riccadonna, S., Merler, S., Chierici, M., Furlanello, C., 2008. Machine learning methods for predictive proteomics. Briefings in bioinformatics 9, 119–128.

Bennett, A., Garner, R., Morris, M.D., La Rocca, M., Barisano, G., Cua, R., Loon, J., Alba, C., Carbone, P., Gao, S., et al., 2023. Manual lesion segmentations for traumatic brain injury characterization. Frontiers in Neuroimaging 2, 14.

Brown, G., 2009. A new perspective for information theoretic feature selection, in: Artificial intelligence and statistics, PMLR. pp. 49–56.

Chang, E.F., Meeker, M., Holland, M.C., 2006. Acute traumatic intraparenchymal hemorrhage: risk factors for progression in the early post-injury period. Neurosurgery 58, 647–656.

Chapman, J., Wang, H.T., 2021. Cca-zoo: A collection of regularized, deep learning based, kernel, and probabilistic cca methods in a scikit-learn style framework. Journal of Open Source Software 6, 3823. URL: 10.21105/joss.03823, doi:10.21105/joss.03823.

Choi, J.H., Lee, J.S., 2019. Embracenet: A robust deep learning architecture for multimodal classification. Information Fusion 51, 259–270.

Crinion, J., Ashburner, J., Leff, A., Brett, M., Price, C., Friston, K., 2007. Spatial normalization of lesioned brains: performance evaluation and impact on fmri analyses. Neuroimage 37, 866–875.

Dash, M., Liu, H., 1997. Feature selection for classification. Intelligent data analysis 1, 131–156.

Dunn, O.J., 1961. Multiple comparisons among means. Journal of the American statistical association 56, 52–64.

Džeroski, S., Ženko, B., 2004. Is combining classifiers with stacking better than selecting the best one? Machine learning 54, 255–273.

Erdogmus, D., Ozertem, U., Lan, T., 2007. Information Theoretic Feature Selection and Projection. volume 83. pp. 1–22. doi:10.1007/978-3-540-75398-81.

Faghihpirayesh, R., Ruf, S., La Rocca, M., Garner, R., Vespa, P., Erdogmus, D., Duncan, D., 2021. Automatic detection of eeg epileptiform abnormalities in traumatic brain injury using deep learning, in: IEEE Engineering in Medicine and Biology Society.

Faria, A.V., Joel, S.E., Zhang, Y., Oishi, K., van Zjil, P.C., Miller, M.I., Pekar, J.J., Mori, S., 2012. Atlas-based analysis of resting-state functional connectivity: Evaluation for reproducibility and multi-modal anatomy–function correlation studies. Neuroimage 61, 613–621.

Feng, Z., Prentice, R., Srivastava, S., 2004. Research issues and strategies for genomic and proteomic biomarker discovery and validation: a statistical perspective. Pharmacogenomics 5, 709–719.

Gugger, J.J., Diaz-Arrastia, R., 2022. Early Posttraumatic Seizures—Putting Things in Perspective. JAMA Neurology 79, 325–326.

Gupta, R.K., Saksena, S., Agarwal, A., Hasan, K.M., Husain, M., Gupta, V., Narayana, P.A., 2005. Diffusion tensor imaging in late posttraumatic epilepsy. Epilepsia 46, 1465–1471.

Hajian-Tilaki, K., 2013. Receiver operating characteristic (roc) curve analysis for medical diagnostic test evaluation. Caspian journal of internal medicine 4, 627.

Hanley, J.A., McNeil, B.J., 1982. The meaning and use of the area under a receiver operating characteristic (roc) curve. Radiology 143, 29–36.

Hotelling, H., 1992. Relations between two sets of variates, in: Breakthroughs in statistics. Springer, pp. 162–190.

Irimia, A., Goh, S.Y.M., Wade, A.C., Patel, K., Vespa, P.M., Van Horn, J.D., 2017. Traumatic brain injury severity, neuropathophysiology, and clinical outcome: insights from multimodal neuroimaging. Frontiers in neurology 8, 530.

Jenkinson, M., Beckmann, C.F., Behrens, T.E., Woolrich, M.W., Smith, S.M., 2012. Fsl. Neuroimage 62, 782–790.

Jie, N.F., Zhu, M.H., Ma, X.Y., Osuch, E.A., Wammes, M., Théberge, J., Li, H.D., Zhang, Y., Jiang, T.Z., Sui, J., et al., 2015. Discriminating bipolar disorder from major depression based on svm-foba: efficient feature selection with multimodal brain imaging data. IEEE transactions on autonomous mental development 7, 320–331.

Kim, J.A., Boyle, E.J., Wu, A.C., Cole, A.J., Staley, K.J., Zafar, S., Cash, S.S., Westover, M.B., 2018. Epileptiform activity in traumatic brain injury predicts post-traumatic epilepsy. Annals of Neurology 83, 858–862.

Kohavi, R., John, G.H., 1997. Wrappers for feature subset selection. Artificial intelligence 97, 273–324.

La Rocca, M., Barisano, G., Garner, R., Ruf, S.F., Amoroso, N., Monti, M., Vespa, P., Bellotti, R., Erdoğmuş, D., Toga, A.W., et al., 2023. Functional connectivity alterations in traumatic brain injury patients with late seizures. Neurobiology of Disease 179, 106053.

Lewis, J.J., O’Callaghan, R.J., Nikolov, S.G., Bull, D.R., Canagarajah, N., 2007. Pixel-and region-based image fusion with complex wavelets. Information fusion 8, 119–130.

Liu, J., Liu, X., Zhang, Y., Zhang, P., Tu, W., Wang, S., Zhou, S., Liang, W., Wang, S., Yang, Y., 2021. Self-representation subspace clustering for incomplete multi-view data, in: Proceedings of the 29th ACM International Conference on Multimedia, pp. 2726–2734.

Lundberg, S.M., Lee, S.I., 2017. A unified approach to interpreting model predictions. Advances in neural information processing systems 30.

Lutkenhoff, E.S., Shrestha, V., Tejeda, J.R., Real, C., McArthur, D.L., Duncan, D., La Rocca, M., Garner, R., Toga, A.W., Vespa, P.M., et al., 2020. Early brain biomarkers of post-traumatic seizures: initial report of the multicentre epilepsy bioinformatics study for antiepileptogenic therapy (epibios4rx) prospective study. Journal of Neurology, Neurosurgery & Psychiatry 91, 1154–1157.

McDermott, J.E., Wang, J., Mitchell, H., Webb-Robertson, B.J., Hafen, R., Ramey, J., Rodland, K.D., 2013. Challenges in biomarker discovery: combining expert insights with statistical analysis of complex omics data. Expert opinion on medical diagnostics 7, 37–51.

Memarian, N., Kim, S., Dewar, S., Engel, J., Staba, R.J., 2015. Multimodal data and machine learning for surgery outcome prediction in complicated cases of mesial temporal lobe epilepsy. Computers in Biology and Medicine 64, 67–78.

Meng, L., Xiang, J., Kotecha, R., Rose, D., Zhao, H., Zhao, D., Yang, J., Degrauw, T., 2010. White matter abnormalities in children and adolescents with temporal lobe epilepsy. Magnetic resonance imaging 28, 1290–1298.

Murphy, K.P., 2012. Machine learning: a probabilistic perspective. MIT press.

Owen, T.W., de Tisi, J., Vos, S.B., Winston, G.P., Duncan, J.S., Wang, Y., Taylor, P.N., 2021. Multivariate white matter alterations are associated with epilepsy duration. European Journal of Neuroscience 53, 2788–2803.

Özdenizci, O., Erdoğmuş, D., 2021. Stochastic mutual information gradient estimation for dimensionality reduction networks. Information Sciences 570, 298–305.

Pedregosa, F., Varoquaux, G., Gramfort, A., Michel, V., Thirion, B., Grisel, O., Blondel, M., Prettenhofer, P., Weiss, R., Dubourg, V., Vanderplas, J., Passos, A., Cournapeau, D., Brucher, M., Perrot, M., Duchesnay, E., 2011. Scikit-learn: Machine learning in Python. Journal of Machine Learning Research 12, 2825–2830.

Piccenna, L., Shears, G., O’Brien, T.J., 2017. Management of post-traumatic epilepsy: An evidence review over the last 5 years and future directions. Epilepsia open 2, 123–144.

Rolls, E.T., Huang, C.C., Lin, C.P., Feng, J., Joliot, M., 2020. Automated anatomical labelling atlas 3. Neuroimage 206, 116189.

Scheffer, J., 2002. Dealing with missing data.

Sharma, A., Garner, R., La Rocca, M., Alba, C., Lee, Y., Yang, K., Brawer-Cohen, M., Duncan, D., 2021. Machine learning of diffusion weighted imaging for prediction of seizure susceptibility following traumatic brain injury, in: 2021 Annual Modeling and Simulation Conference (ANNSIM), IEEE. pp. 1–9.

Sharp, D.J., Beckmann, C.F., Greenwood, R., Kinnunen, K.M., Bonnelle, V., De Boissezon, X., Powell, J.H., Counsell, S.J., Patel, M.C., Leech, R., 2011. Default mode network functional and structural connectivity after traumatic brain injury. Brain 134, 2233–2247.

Singh, A., Westlin, C., Eisenbarth, H., Reynolds Losin, E.A., Andrews-Hanna, J.R., Wager, T.D., Satpute, A.B., Barrett, L.F., Brooks, D.H., Erdogmus, D., 2021. Variation is the norm: Brain state dynamics evoked by emotional video clips, in: 2021 43rd Annual International Conference of the IEEE Engineering in Medicine and Biology Society (EMBC), pp. 6003–6007.

Smith, S.M., Jenkinson, M., Johansen-Berg, H., Rueckert, D., Nichols, T.E., Mackay, C.E., Watkins, K.E., Ciccarelli, O., Cader, M.Z., Matthews, P.M., et al., 2006. Tract-based spatial statistics: voxelwise analysis of multi-subject diffusion data. Neuroimage 31, 1487–1505.

Sourati, J., Erdogmus, D., Akcakaya, M., Kazmierczak, S.C., Leen, T.K., 2015. A NOVEL DELTA CHECK METHOD FOR DETECTING LABORATORY ERRORS Northeastern University University of Pittsburgh Oregon Health & Science University Portland, OR, USA National Science Foundation, 0–5.

Temkin, N.R., 2009. Preventing and treating posttraumatic seizures: the human experience. Epilepsia 50, 10–13.

Tenenhaus, A., Tenenhaus, M., 2011. Regularized generalized canonical correlation analysis. Psychometrika 76, 257–284.

Troyanskaya, O., Cantor, M., Sherlock, G., Brown, P., Hastie, T., Tibshirani, R., Botstein, D., Altman, R.B., 2001. Missing value estimation methods for dna microarrays. Bioinformatics 17, 520–525.

Vespa, P., Tubi, M., Claassen, J., Buitrago-Blanco, M., McArthur, D., Velazquez, A.G., Tu, B., Prins, M., Nuwer, M., 2016. Metabolic crisis occurs with seizures and periodic discharges after brain trauma. Annals of Neurology 79, 579–590.

Vespa, P.M., Shrestha, V., Abend, N., Agoston, D., Au, A., Bell, M.J., Bleck, T.P., Blanco, M.B., Claassen, J., Diaz-Arrastia, R., et al., 2019. The epilepsy bioinformatics study for anti-epileptogenic therapy (epibios4rx) clinical biomarker: study design and protocol. Neurobiology of disease 123, 110–114.

Whitfield-Gabrieli, S., Nieto-Castanon, A., 2012. Conn: a functional connectivity toolbox for correlated and anticorrelated brain networks. Brain connectivity 2, 125–141.

Williams, P.L., Beer, R.D., 2010. Nonnegative decomposition of multivariate information. arXiv preprint arXiv:1004.2515 .

Wolpert, D.H., 1992. Stacked generalization. Neural networks 5, 241–259.

Yang, Y., Truong, N.D., Eshraghian, J.K., Maher, C., Nikpour, A., Kavehei, O., 2022. A multimodal ai system for out-of-distribution generalization of seizure identification. IEEE Journal of Biomedical and Health Informatics .

Yushkevich, P.A., Piven, J., Hazlett, H.C., Smith, R.G., Ho, S., Gee, J.C., Gerig, G., 2006. User-guided 3d active contour segmentation of anatomical structures: significantly improved efficiency and reliability. Neuroimage 31, 1116–1128.

Zhang, Y., Zhang, J., Oishi, K., Faria, A.V., Jiang, H., Li, X., Akhter, K., Rosa-Neto, P., Pike, G.B., Evans, A., Toga, A.W., Woods, R., Mazziotta, J.C., Miller, M.I., van Zijl, P.C., Mori, S., 2010. Atlas-guided tract reconstruction for automated and comprehensive examination of the white matter anatomy. NeuroImage 52, 1289–1301.

Zhang, Y.D., Dong, Z., Wang, S.H., Yu, X., Yao, X., Zhou, Q., Hu, H., Li, M., Jiménez-Mesa, C., Ramirez, J., et al., 2020. Advances in multimodal data fusion in neuroimaging: overview, challenges, and novel orientation. Information Fusion 64, 149–187.

